# Applying health equity implementation science frameworks to population genetic screening

**DOI:** 10.1101/2024.03.17.24304021

**Authors:** Nandana D. Rao, Stephanie M. Fullerton, Brian H. Shirts, Annie T. Chen, Nora B. Henrikson

## Abstract

Implementation science frameworks with a focus on health equity have emerged to help guide the introduction of new interventions into healthcare and community settings while limiting health disparities. The purpose of this research was to explore the applicability of such frameworks to guide the equitable implementation of population genetic screening programs. We searched PubMed and reference lists for relevant frameworks and examples of their use in health settings. We then assessed if and how selected frameworks provide guidance for different stages of population genetic screening: recruitment, sample collection, result return, follow-up care and prevention, and cascade screening. Findings were synthesized into a list of health equity considerations specific to each stage. We identified 5 implementation frameworks that focus on health equity. Guidance varied by framework type: determinant (explaining what affects implementation outcomes), process (translating research into practice), or evaluation (assessing implementation). Common characteristics included focusing implementation efforts on populations who have historically experienced health inequities and adapting interventions to fit local contexts. Process models also highlighted the importance of community partnerships. Overall, frameworks offered broad recommendations applicable to population genetic screening program implementation. However, gaps still exist in guidance provided for later stages of population genetic screening. To improve the equitable implementation of genetic screening, future programs may benefit from utilizing one or more of these frameworks or by incorporating the health equity considerations and outcomes compiled in this analysis.

## INTRODUCTION

Population genetic screening, or genetic screening of people regardless of personal or family history of disease, has been proposed to increase the reach of genetic services and identify more people at risk for preventable conditions (Grzymski et al., 2020; King et al., 2014; Limburg et al., 2011). However, population screening is complex for many reasons including the need to appropriately inform large numbers of people about the benefits and harms of genetic screening, collecting DNA samples, and following people over time to ensure they receive appropriate care based on their results. If not implemented with care and the needs of underserved people in mind, population genetic screening may perpetuate or further exacerbate already existing health disparities (Grzymski et al., 2018).

To limit harmful consequences, health equity must be a central consideration in the design and implementation of population genetic screening programs. Health equity is defined as everyone having a fair and just opportunity to be as healthy as possible (Centers for Disease Control and Prevention, n.d.). Striving for health equity requires focusing on the needs of those who are at greatest risk of poor health due to social circumstances (Braveman, 2014). It involves the elimination of health differences that are linked to social determinants historically connected to exclusion, such as race, ethnicity, socioeconomic status, gender, age, religion, disability, sexual orientation, gender identity, and geographic location (Braveman, 2014).

Implementation science frameworks with a focus on health equity have emerged to guide the introduction of new interventions into healthcare and community settings, and their principles could improve the incorporation of genomic discoveries into healthcare (Chambers et al., 2016; Roberts et al., 2017). How well such frameworks suit population genomic screening programs is not well understood.

We conducted a systematic literature search to identify and describe published equity-focused implementation science frameworks and assessed the applicability of these frameworks to population genetic screening programs.

## METHODS

### Population genetic screening stages

We conceptualized population genetic screening in 5 major stages (Figure 1) based on the design of existing pilot screening programs (David et al., 2021; East et al., 2021; Grzymski et al., 2018; Rao et al., 2023). Our descriptive model’s stages include recruitment, sample collection, return of results, follow-up care and prevention, and cascade screening (the notification of biological relatives about genetic risk). This model formed the basis for our analysis of framework applicability to population screening.

**Figure 1.**
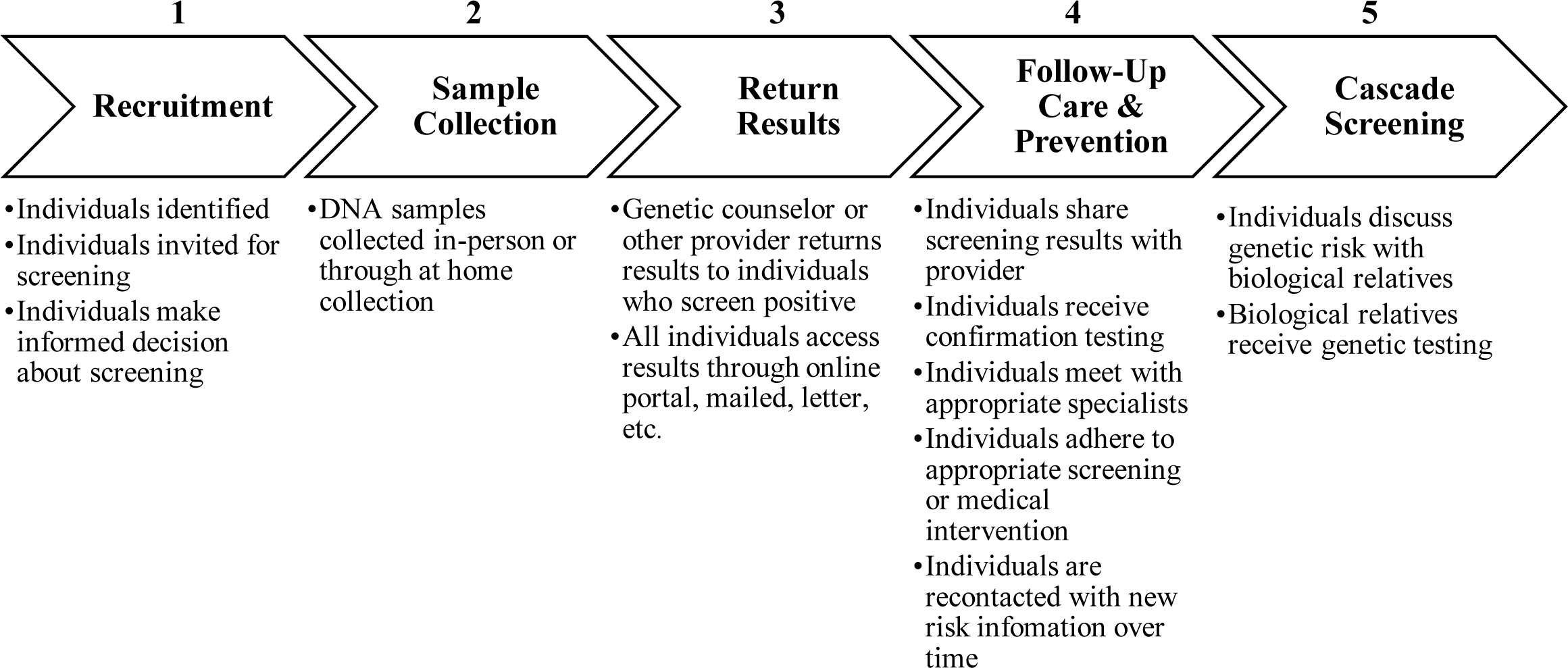
Five major stages of population genetic screening.

### Framework identification

We searched PubMed for frameworks designed to promote health equity during the implementation of health interventions using the following keywords: (“health equity” or “health disparities” or “health inequalities”) and (“implementation” or “translation”) and (“framework” or “model” or “theory”). One author, NDR, screened the resulting titles and abstracts. When articles cited potentially relevant frameworks or included a review of frameworks, NDR examined the reference lists for pertinent publications. Other frameworks previously known to the authors were also considered. Article review was restricted to work published between January 2010 and December 2021, as the focus on health equity in implementation science has become more prominent relatively recently (Odeny, 2021).

Criteria for inclusion were frameworks focused on health equity and implementation of health services, and that were developed for high-resource settings, as these are the most relevant to population genetic screening. Frameworks were excluded if they were specific to a certain health condition or intervention, provided little guidance for implementation or if the article was not available in English. Discussion papers, or those that only described a need for considering health equity during program implementation or provided no explicit framework or model, were also excluded.

### Data extraction and evaluation

For each of the selected frameworks, the following data was extracted: name, author, year of publication, type, audience, development, and description. Framework type was determined according to Nilsen’s categorizations of implementation science theories, models, and frameworks: determinant frameworks, process models, or evaluation frameworks (Nilsen, 2015). Determinant frameworks are designed to assist with understanding barriers or facilitators that influence implementation outcomes; process models to guide the process of translating research into practice; and evaluation frameworks to specify implementation outcomes (Nilsen, 2015).

### Data synthesis

We assessed the applicability of each framework to population genetic screening by evaluating if and how the framework provided guidance for the 5 major stages of population genetic screening we identified. To further our understanding of potential framework application, we looked for examples of how each framework may have been used in other settings by examining articles published through December 2021 that cited our selected frameworks. We also searched for empiric evidence of framework validation.

We then compared the selected frameworks and discussed their strengths and weaknesses with respect to guiding the implementation of population genetic screening. Using findings from our applicability assessment, we also compiled a list of health equity considerations and outcomes specific to each stage of population genetic screening.

## RESULTS

The initial PubMed search yielded 1,013 results. An additional 37 articles were identified through reference lists or because they were previously known to authors. Records were screened by title and abstract followed by full text review (Figure 2). We identified five frameworks designed to reduce or prevent health disparities during the implementation of health interventions (Table 1). One framework was described in two of the selected articles.

**Figure 2.**
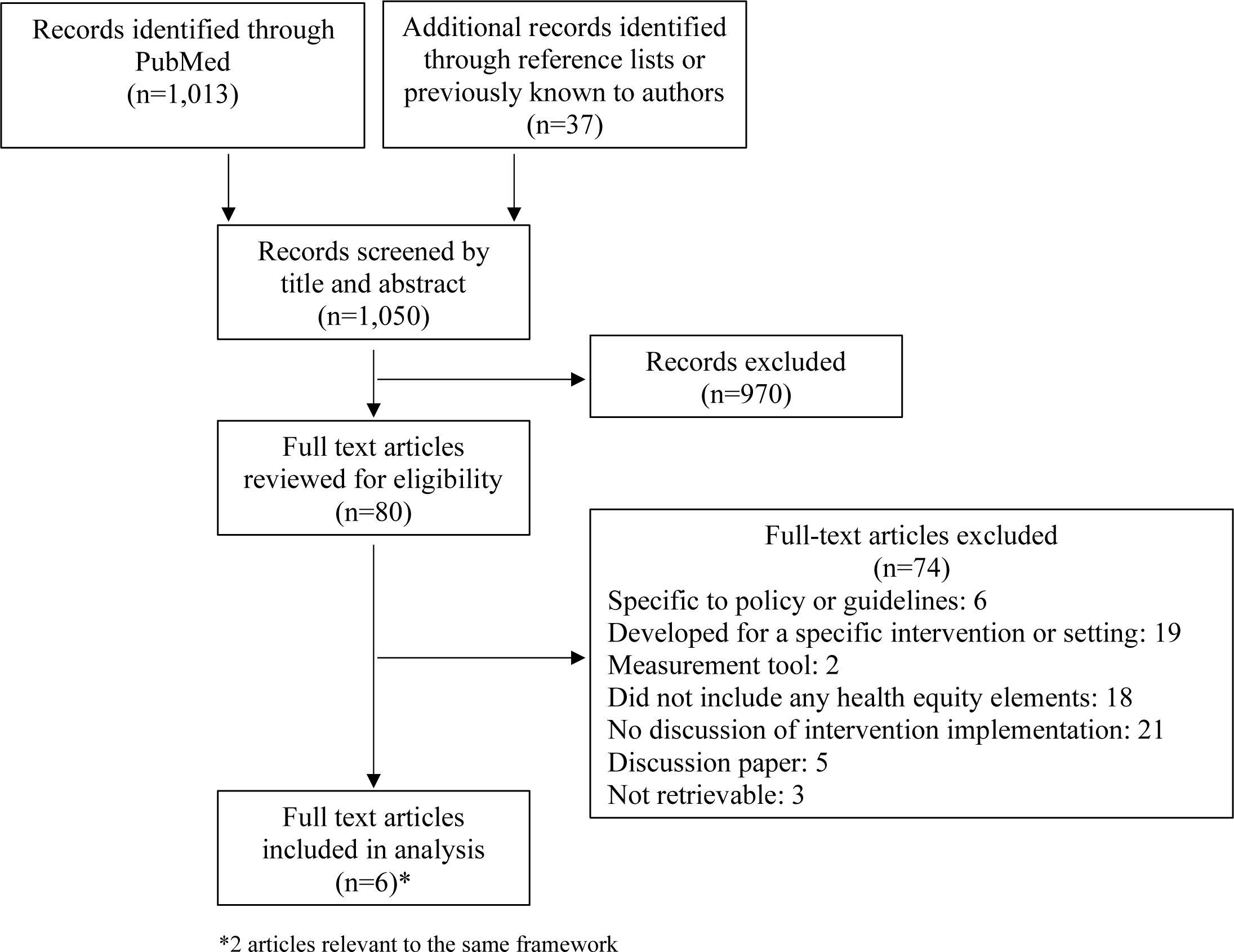
Diagram of article search and selection process.

**Table 1:**
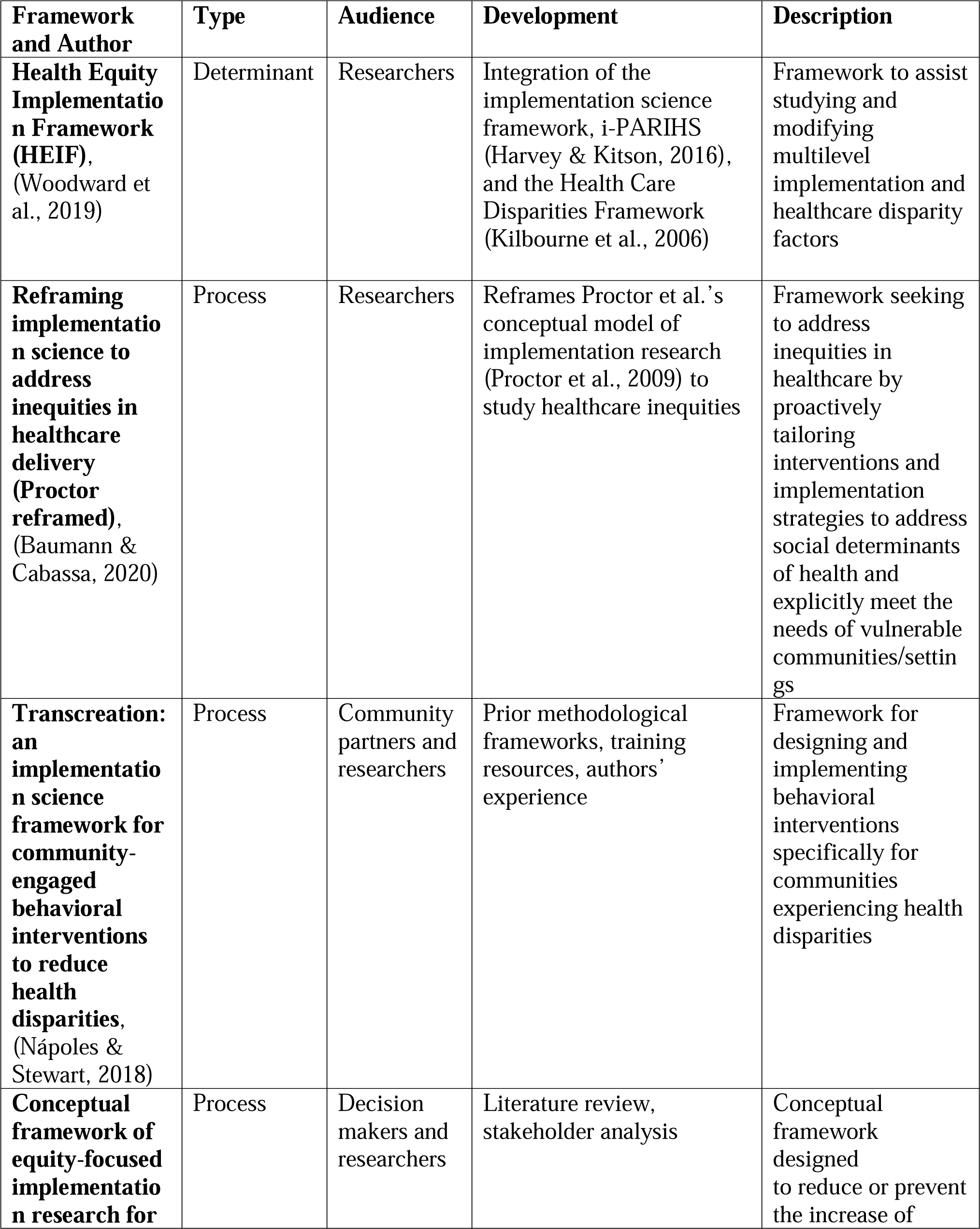
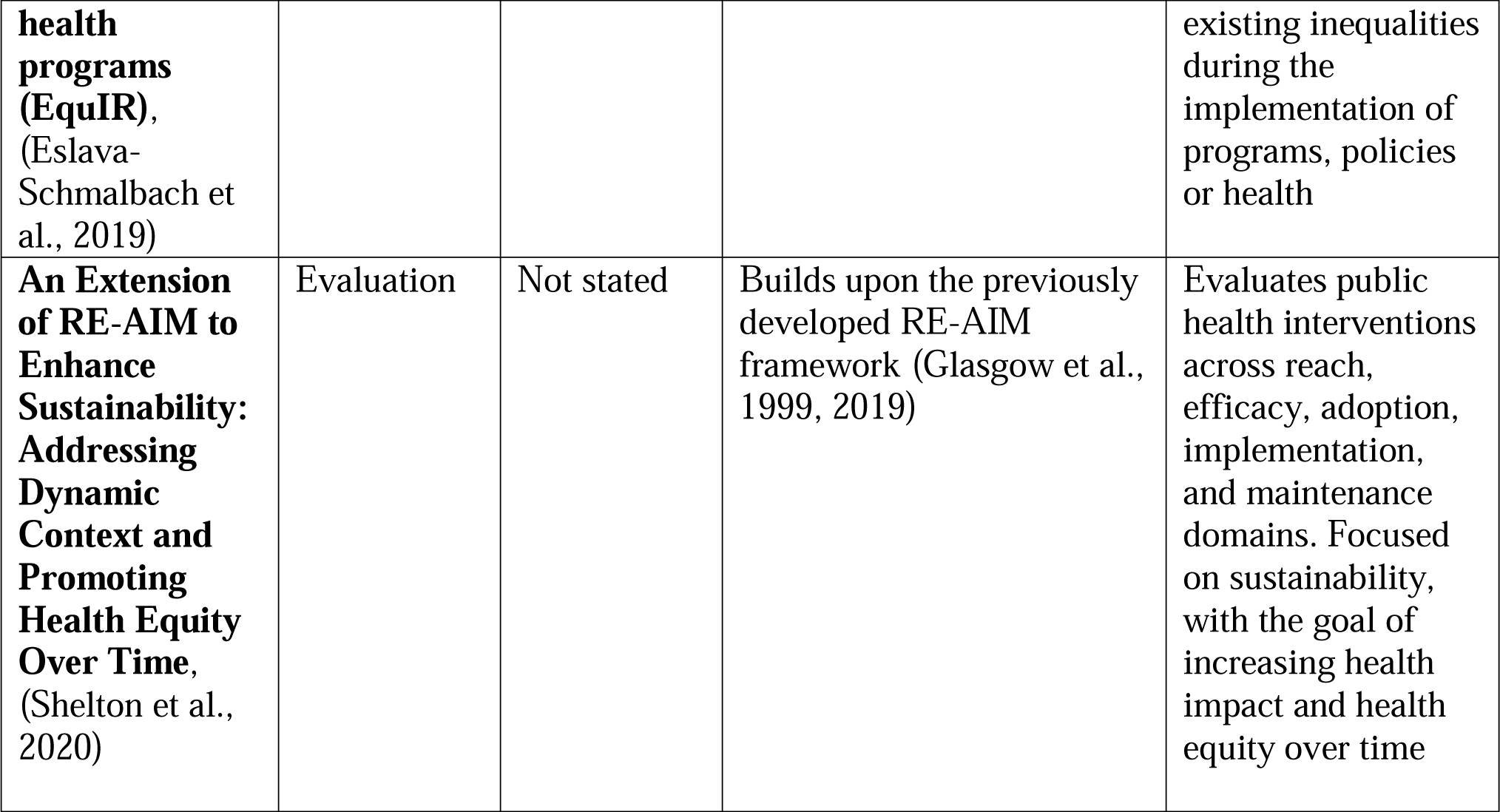
Characteristics of included frameworks.

| **Framework and Author** | **Type** | **Audience** | **Development** | **Description** |
| --- | --- | --- | --- | --- |
| **Health Equity Implementation Framework (HEIF)**,<br>(Woodward et al., 2019) | Determinant | Researchers | Integration of the implementation science framework, i-PARIHS (Harvey & Kitson, 2016), and the Health Care Disparities Framework (Kilbourne et al., 2006) | Framework to assist studying and modifying multilevel implementation and healthcare disparity factors |
| **Reframing implementation science to address inequities in healthcare delivery (Proctor reframed)**,<br>(Baumann & Cabassa, 2020) | Process | Researchers | Reframes Proctor et al.'s conceptual model of implementation research (Proctor et al., 2009) to study healthcare inequities | Framework seeking to address inequities in healthcare by proactively tailoring interventions and implementation strategies to address social determinants of health and explicitly meet the needs of vulnerable communities/settings |
| **Transcreation: an implementation science framework for community-engaged behavioral interventions to reduce health disparities**,<br>(Nápoles & Stewart, 2018) | Process | Community partners and researchers | Prior methodological frameworks, training resources, authors' experience | Framework for designing and implementing behavioral interventions specifically for communities experiencing health disparities |
| **Conceptual framework of equity-focused implementation research for** | Process | Decision makers and researchers | Literature review, stakeholder analysis | Conceptual framework designed to reduce or prevent the increase of |
| **health programs (EquIR),**<br>(Eslava-Schmalbach et al., 2019) |  |  |  | existing inequalities during the implementation of programs, policies or health |
| **An Extension of RE-AIM to Enhance Sustainability: Addressing Dynamic Context and Promoting Health Equity Over Time,**<br>(Shelton et al., 2020) | Evaluation | Not stated | Builds upon the previously developed RE-AIM framework (Glasgow et al., 1999, 2019) | Evaluates public health interventions across reach, efficacy, adoption, implementation, and maintenance domains. Focused on sustainability, with the goal of increasing health impact and health equity over time |

We categorized the frameworks as one determinant framework (HEIF, Woodward et al., 2019), three process frameworks (Proctor reframed, Baumann & Cabassa, 2020; Transcreation, Nápoles & Stewart, 2018; EquIR, Eslava-Schmalbach et al., 2019), and one evaluation framework (RE-AIM extension, Shelton et al., 2020). Researchers were the primary intended audience for each framework. Frameworks were largely conceptually developed by the authors, though one, EquIR (Eslava-Schmalbach et al., 2019), was developed using stakeholder engagement (Table 2). Our search for evidence of framework validation yielded no information about if and how any of the five frameworks had been validated.

**Table 2:** Components of included frameworks.

| Framework | Components/Steps |
| --- | --- |
| **HEIF**<br>(Woodward et al., 2019) | Factors to understand healthcare disparity determinants:<br>• Clinical encounter: patient-provider interaction<br>• Culturally relevant factors: characteristics unique to a group of people in the implementation effort based on their lived experience<br>• Societal context: physical structures, economies, sociopolitical forces<br>• Context: micro, meso, or macro levels that correspond to inner and outer contexts<br>• Recipients: individuals who influence implementation and those who are affected by its outcomes<br>• Innovation: characteristics of the treatment, intervention, practice, or new “thing” to be implemented<br>• Facilitation: implementation strategies that result in implementation coming to fruition |
| **Proctor reframed**<br>(Baumann & Cabassa, 2020) | Steps to design intervention and implementation strategies to address healthcare inequities:<br>1. Focus on reach from the very beginning<br>2. Design and select interventions for vulnerable populations with implementation in mind<br>3. Implement what works and develop implementation strategies that can help reduce inequities in care<br>4. Develop the science of adaptation<br>5. Use an equity lens for implementation outcomes |
| **Transcreation**<br>(Nápoles & Stewart, 2018) | Steps involved in designing, delivering, and evaluating intervention to reduce health disparities:<br>1. Identify community infrastructure and engage partners<br>2. Specify theory<br>3. Identify multiple inputs for new program<br>4. Design intervention prototype<br>5. Design study, methods, and measure for community setting<br>6. Build community capacity for delivery<br>7. Deliver “transcreated” intervention (e.g., an intervention designed to resonate with the intended community and reduce health disparities) and evaluate implementation processes |
| **EquIR**<br>(Eslava-Schmalbach et al., 2019) | Cyclical steps to prevent the increase of inequalities during intervention implementation:<br>1. Identify the health status of the population, including potentially disadvantaged population(s)<br>2. Identify relevant research questions given the disadvantaged populations, quantify the inequalities to be solved, develop equity-sensitive recommendations for implementation<br>3. Identify key players and barriers and facilitators for the implementation of equity-sensitive recommendations<br>4. Design strategies to overcome identified barriers, define monitoring and evaluation strategies, and design the equity-focused communication strategies<br>5. Monitor implementation outcomes using an equity focus (outcomes listed below)<br>6. Return to step 1 – Population health status after implementation is the new starting point for further implementation<br>Implementation outcomes to evaluate equity:<br>• Acceptability: perception among key implementation players including health professionals, stakeholders, patients, community, disadvantaged population<br>• Adoption: intention, utilization, or action to try to employ the sensitive equity recommendation in the new program or intervention<br>• Appropriateness: relevance or perceived fit, or usefulness or practicability of the program or intervention in the disadvantaged population<br>• Feasibility: extent to which the program or intervention allows to reduce the barriers, and can be carried out in any setting, especially among disadvantaged populations<br>• Fidelity: adherence of disadvantaged population to the equity-focused implementation program or intervention<br>• Implementation cost: Total cost of the program implementation in disadvantaged and non-disadvantaged populations, and the final adjusted cost- effectiveness economic evaluation<br>• Coverage: degree of reach, access, service spread or effective coverage (combining coverage and fidelity) on the disadvantaged population eligible to benefit from the program or the intervention<br>• Sustainability: maintenance, continuation or durability of the program or |
|  | intervention implemented through short, medium and long- term strategies, including disadvantaged populations |
| **RE-AIM extension**<br>(Shelton et al., 2020) | Health equity considerations for evaluation domains:<br>• Reach: Considering social determinants of health (SDOH), who is reached by intervention and who is not? Why? How can reach be improved for populations who are experiencing inequities?<br>• Effectiveness: Are health impacts equitable across all groups based on SDOH? Why or why not? Do certain populations experience higher levels of negative effects?<br>• Adoption: Did all settings adopt the intervention equitably? Which settings staff did/did not and why? Were low-resource settings able to adopt the intervention to the same extent as higher-resource settings? What adaptations will facilitate adoption?<br>• Implementation: Were the intervention and implementation strategies equitably delivered across settings/staff? Which settings/staff were/were not successful in delivery and why? Do all settings/staff have capacity/resources to deliver the intervention on an ongoing basis? What adaptations are needed to promote equity and address SDOH?<br>• Maintenance: Is the intervention being equitably sustained? What settings/populations continue to be reached by the intervention over time? Why? Do intervention adaptations exacerbate inequities over time? Do all settings have capacity to maintain delivery of the intervention? Are sustainability determinants the same across low and high-resource settings? How do SDOH impact inequitable implementation and sustainability? |

### The Health Equity Implementation Framework (HEIF)

#### Description

Woodward and colleagues developed HEIF by integrating the i-PARIHS implementation science framework (Harvey & Kitson, 2016) and the Health Care Disparities Framework (Kilbourne et al., 2006). HEIF is designed to help researchers determine factors related to innovation uptake and disparities in healthcare to improve outcomes for marginalized populations (Woodward et al., 2019). Health equity domains include culturally relevant factors, the clinical encounter, and societal context (Table 2). Culturally relevant factors are specific to intervention recipients based on their lived experience and can include characteristics such as socioeconomic status, implicit bias, health literacy, trust in providers, language, race and ethnicity. The clinical encounter encompasses interactions between providers and patients, which influence if an intervention is offered by a provider or accepted by a patient. These encounters are influenced by inner context at the local (e.g., clinic) and organizational (e.g., hospital) levels, and outer context (e.g., the healthcare system). Finally, the societal context includes economies, physical structures (how environments are built or arranged), and sociopolitical forces (social norms or political forces). These impact health disparities by influencing the inner and outer context, the clinical encounter, and culturally relevant factors. The HEIF has previously been applied to design an interview guide and direct content analysis to identify implementation factors and best practices for social needs screening in primary care settings (Drake et al., 2021).

#### Application to population genetic screening

The HEIF is well suited to provide guidance for anticipating possible barriers or facilitators to implementation across all stages of population genetic screening (Table 3). For example, during recruitment, attention to cultural factors can help researchers anticipate how language and cultural beliefs influence informed consent and enrollment. HEIF’s physical structures domain can inform how in-person sample collection and return of results may facilitate or impede screening depending on access to reliable transportation and the location of facilities. Similarly, during follow-up care and prevention, understanding potentially inequitable physical spaces can inform implementation. In the cascade screening stage, reflecting on sociopolitical forces, such genetic privacy laws, may illuminate barriers to information sharing among relatives.

**Table 3:**
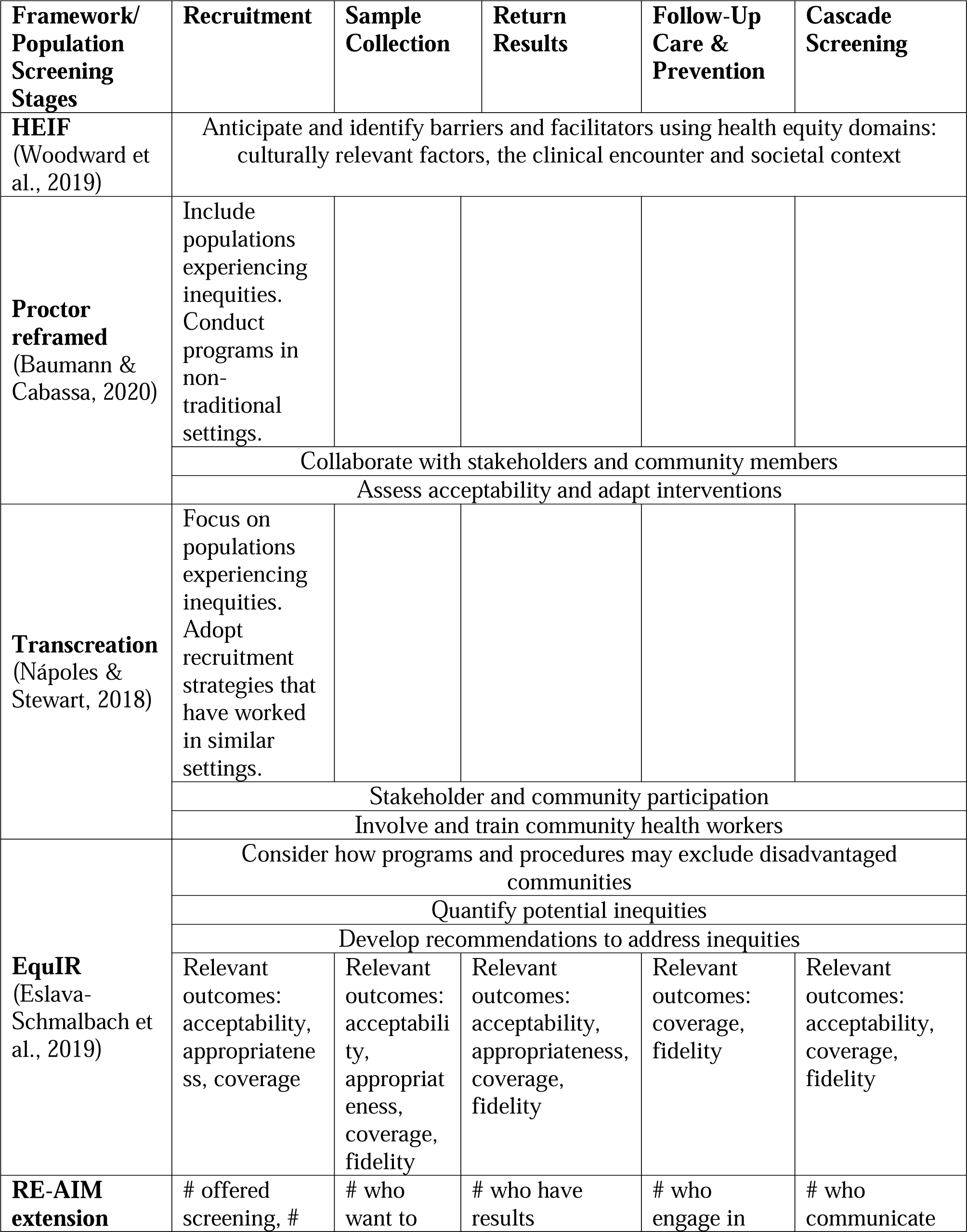
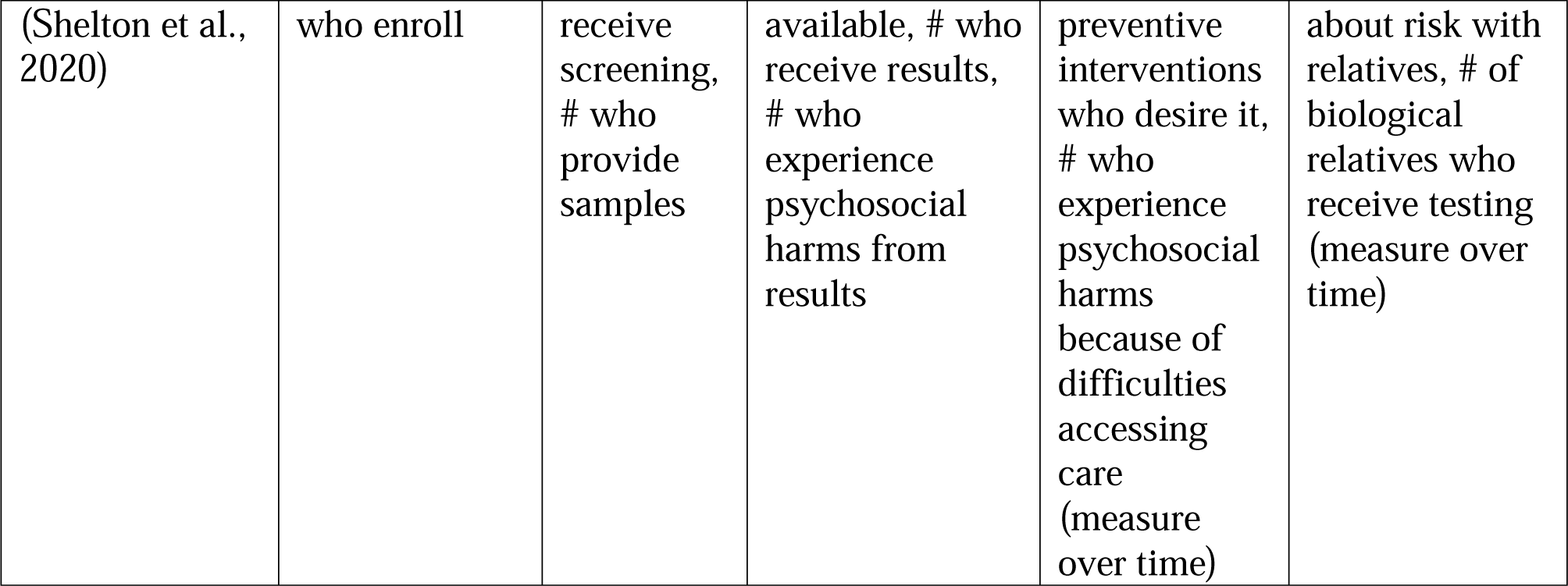
Applicability of frameworks to population genetic screening programs.

| **Framework/**<br>**Population**<br>**Screening**<br>**Stages** | **Recruitment** | **Sample**<br>**Collection** | **Return**<br>**Results** | **Follow-Up**<br>**Care &**<br>**Prevention** | **Cascade**<br>**Screening** |
| --- | --- | --- | --- | --- | --- |
| **HEIF**<br>(Woodward et al., 2019) | Anticipate and identify barriers and facilitators using health equity domains: culturally relevant factors, the clinical encounter and societal context |  |  |  |  |
| **Proctor**<br>**reframed**<br>(Baumann & Cabassa, 2020) | Include populations experiencing inequities. Conduct programs in non-traditional settings. |  |  |  |  |
|  | Collaborate with stakeholders and community members |  |  |  |  |
|  | Assess acceptability and adapt interventions |  |  |  |  |
| **Transcreation**<br>(Nápoles & Stewart, 2018) | Focus on populations experiencing inequities. Adopt recruitment strategies that have worked in similar settings. |  |  |  |  |
|  | Stakeholder and community participation |  |  |  |  |
|  | Involve and train community health workers |  |  |  |  |
| **EquiR**<br>(Eslava-Schmalbach et al., 2019) | Consider how programs and procedures may exclude disadvantaged communities |  |  |  |  |
|  | Quantify potential inequities |  |  |  |  |
|  | Develop recommendations to address inequities |  |  |  |  |
|  | Relevant outcomes: acceptability, appropriateness, coverage | Relevant outcomes: acceptability, appropriateness, coverage, fidelity | Relevant outcomes: acceptability, appropriateness, coverage, fidelity | Relevant outcomes: coverage, fidelity | Relevant outcomes: acceptability, coverage, fidelity |
| **RE-AIM**<br>**extension** | # offered screening, # | # who want to | # who have results | # who engage in | # who communicate |
| (Shelton et al., 2020) | who enroll | receive screening, # who provide samples | available, # who receive results, # who experience psychosocial harms from results | preventive interventions who desire it, # who experience psychosocial harms because of difficulties accessing care (measure over time) | about risk with relatives, # of biological relatives who receive testing (measure over time) |

### Proctor reframed

#### Description

Baumann and Cabassa (2020) reframed the Proctor implementation science framework to provide an example of how to apply an existing framework to address healthcare inequities. The original Proctor framework posits that interventions differ from their implementation strategies and requires the involvement of various stakeholders at multiple levels (Proctor et al., 2009). The original Proctor also proposes outcomes in three interrelated but distinct domains: implementation (e.g., feasibility, fidelity, acceptance), service (e.g., efficiency, safety, effectiveness), and client (e.g., satisfaction, function). The reframed Proctor framework emphasizes collaborating with stakeholders and community members throughout intervention planning, design, and implementation in order to understand and meet the needs of historically underserved communities (Table 2) (Baumann & Cabassa, 2020). It proposes continually adapting programs based on the needs of populations with the goal of reducing inequities through systematic changes to intervention and implementation strategies. Finally, Proctor reframed suggests conducting descriptive and explanatory studies to identify factors that contribute to inequities in implementation outcomes. We did not identify any published applications of Proctor-reframed in health settings.

#### Application to population genetic screening

Proctor-reframed specifies guidance most relevant to the recruitment stage, including ensuring that populations that have previously experienced inequities in genetic services are included in population genetic screening programs. Suggested strategies for enhancing inclusion are conducting programs in non-traditional settings such as in faith communities or community centers. This framework also discusses how face-to-face presentations with community members and person-to-person recruitment can assist with enrolling people who would otherwise not participate.

### Transcreation

#### Description

Transcreation is defined as the process of planning and delivering interventions to reduce health disparities that resonate with the intended community (Nápoles & Stewart, 2018). Nápoles and colleagues created this framework to address the differences that occur between original intervention implementation settings (often among mainstream populations or in academic settings) and when interventions are adopted among a population facing health disparities.

Collaboration is a central principle of Transcreation, which proposes stakeholder and community involvement through the entire process of intervention design, implementation, and adaptation (Table 2) (Nápoles & Stewart, 2018). This framework assumes the presence of an established partnership between researchers and community members and a shared understanding of the disparity to be addressed. As part of the framework’s proposed collaboration, Transcreation suggests involving community workers in implementation by training them in intervention delivery. Fitting interventions to context and population needs is also prominent.

Transcreation has previously been applied in other health settings; for example, it has been used to adapt a stress management intervention for Latina breast cancer survivors living in rural settings (Santoyo-Olsson et al., 2019).

#### Application to population genetic screening

Transcreation provides guidance most relevant for the initial recruitment stage of population genetic screening by suggesting focusing attention on populations who experience disparities in access to and utilization of genetic services (Table 3). Through the incorporation of scientific evidence, programs can also adopt recruitment strategies that have been proven to work in similar settings.

Unique to Transcreation is training community members in intervention delivery. This idea is relevant for all population screening stages, as members can be trained to provide cultural, informational and logistical support to specific communities within a general population. During recruitment, this can promote informed decision-making. In the follow-up and prevention stage, community members acting as patient navigators can assist individuals with information about insurance or recommended medical interventions.

### EquIR

#### Description

Eslava-Schmalbach and colleagues developed EquIR for researchers and decision makers to reduce or prevent health inequities during the implementation of health programs or policies (Eslava-Schmalbach et al., 2019). This conceptual framework is cyclic, with social determinants of health considered throughout. The cycle begins with identifying disadvantaged populations and quantifying current health inequalities (Table 2). It then suggests developing and implementing recommendations to meet the needs of disadvantaged populations with key players such as health professionals, patients, community members, and stakeholders. It finishes by recommending the monitoring of implementation outcomes (Table 2) and identifying how the intervention has impacted the health status of populations receiving the intervention. From here the cycle continues and the new population health status becomes the starting point of the intervention. Beyond its initial development, we identified no examples of the use of EquIR to guide implementation of health interventions.

#### Application to population genetic screening

Of the guidance proposed by EquIR, the described outcomes are most readily applied to population genetic screening and can be used to understand how programs impact disadvantaged populations at each stage. For example, measures of acceptability and appropriateness can be applied during recruitment, sample collection, and return of results stages to understand stakeholder perceptions of fit and usefulness of program procedures (Table 3). Measures of fidelity and coverage may be useful during follow-up care and cascade screening stages for understanding how often people receiving positive screening results are able to act on these results and how often risk information is shared with biological relatives. In addition, the cyclical nature of EquIR promotes ongoing program adjustments informed by these outcome measures.

### RE-AIM extension

#### Description

The extension to the RE-AIM framework authored by Shelton and colleagues is designed to promote sustainability and health equity. The original RE-AIM framework focuses on evaluation and includes both individual and staff/setting level domains: Reach and effectiveness (individual), adoption and implementation (staff/setting), and maintenance (individual and staff/setting) (Glasgow et al., 1999, 2019). While the extension to RE-AIM discusses these same domains and previously described indicators, Shelton et al. provide additional guidance to consider health equity during the measurement of these indicators (Table 2). This guidance focuses on assessing indicators over time across different populations of focus (defined by age, race, ethnicity, disability, insurance status, literacy level or other social determinants of health), to identify and address health inequities (Shelton et al., 2020). The extension to RE-AIM also considers the link between health equity and costs or resources and suggests incorporating cost estimates and resource requirements into planning discussions with stakeholders. This framework has previously been used to evaluate the implementation of a COVID-19 vaccine program seeking to facilitate equitable vaccine access and uptake among Latinx community members (Marquez et al., 2021).

#### Application to population genetic screening

The outcome indicators and health equity considerations listed by RE-AIM extension give measures that can be monitored at each stage of population genetic screening (Table 3). During recruitment, relevant indicators include the number of people who are offered screening and the number of people who agree to screening. Taking into account social determinants of health when interpreting these indicators can determine if all populations are offered and enroll in screening similarly and reveal which populations are not reached. Reach can also be ascertained for sample collection, return of results, and cascade screening to find inequities that may be emerging during these stages.

Measures of effectiveness across social determinants of health are also relevant for the return of results, follow-up care, and cascade screening stages. For return of results, indicators include the number of people experiencing psychosocial harms upon learning results. For follow-up care and prevention, relevant indicators are the number of people who are able to engage in preventive interventions who desire it and the number of people who experience psychosocial harms because of difficulties accessing care. During cascade screening, indicators include the number of biological relatives who receive testing.

### Comparing frameworks

A number of characteristics were shared across the analyzed frameworks. The first was to focus implementation efforts on populations who have historically experienced health inequities. This is a crucial consideration as placing specific emphasis on underserved populations at the beginning of implementation planning can reorient design and procedures to better prioritize the needs of such communities. Another common element across frameworks was to adapt interventions to fit local context and meet the needs of marginalized communities. Doing so can limit the implementation gap, which occurs when the context where interventions are designed and developed does not align with realities of implementation settings. Constraining this gap can increase the appropriateness of an intervention (Baumann & Cabassa, 2020).

The identified frameworks were conceptually focused, rather than validated theories, and the guidance provided varied by framework type, as expected. The process models, Proctor reframed (Baumann & Cabassa, 2020), Transcreation (Nápoles & Stewart, 2018), and EquIR (Eslava-Schmalbach et al., 2019), for example, tended to be high-level, and provided overarching considerations and recommendations for program design, implementation, and evaluation rather than specific guidance that lends itself to individual stages of an intervention like population genetic screening. Regarding screening, Proctor reframed (Baumann & Cabassa, 2020) and Transcreation (Nápoles & Stewart, 2018) recommendations applied most directly to the recruitment stage. While all three process models described evaluating implementation outcomes keeping social determinants and differences in outcomes across populations in mind, they varied in the specificity with which they described and defined these outcomes.

In contrast, the determinant framework, HEIF (Woodward et al., 2019), provides an explicit means to identify barriers to program implementation throughout all stages of population genetic screening. Similarly, the evaluation framework, RE-AIM extension (Shelton et al., 2020), detailed indicators and health equity considerations for monitoring program outcomes relevant to all program stages.

Among the process models, Proctor reframed (Baumann & Cabassa, 2020), Transcreation (Nápoles & Stewart, 2018), and EquIR (Eslava-Schmalbach et al., 2019), another main concept was the importance of involving community partners and other stakeholders throughout implementation. Such collaboration allows researchers to learn more about local customs and build trust with community members (Cabassa & Baumann, 2013; Wallerstein & Duran, 2010). Interventions can better be tailored to a specific population and integrate relevant perspectives, norms, and social and cultural values. As a result, this may improve intervention acceptability and effectiveness and prevent health disparities from emerging.

## DISCUSSION

In this analysis we outline relevant equity considerations for population genetic screening program implementation guided by five selected frameworks: HEIF (Woodward et al., 2019), Proctor reframed (Baumann & Cabassa, 2020), Transcreation (Nápoles & Stewart, 2018), EquIR (Eslava-Schmalbach et al., 2019), and RE-AIM extension (Shelton et al., 2020). The HEIF (Woodward et al., 2019), RE-AIM extension (Shelton et al., 2020), and outcome measures provided in EquIR (Eslava-Schmalbach et al., 2019) were applicable to all stages of population screening. Remaining guidance from EquIR (Eslava-Schmalbach et al., 2019) and ideas proposed by Proctor reframed (Baumann & Cabassa, 2020) and Transcreation (Nápoles & Stewart, 2018) tended to be broad and less applicable to each individual screening stage.

Results of our analysis may offer insights for researchers designing new population genetic screening programs and assist with identification and selection of relevant frameworks to direct implementation. To make the best use of the variety of recommendations brought up by the different frameworks, these frameworks may best be used in tandem. For instance, determinant domains can be used when process model steps suggest identifying implementation barriers and specific indicators can be drawn from evaluation frameworks when steps call for assessing implementation outcomes.

We found that guidance for later stages of population genetic screening programs, such as follow-up care and cascade screening, was limited beyond central framework characteristics. For true public health impact, individuals receiving positive screening results must have access to services to delay or prevent disease onset. Health benefits may also be seen if genetic risk information is communicated to relatives. However, frameworks lacked specific guidance about how to ensure equitable referrals to follow-up care or promote adherence to recommended medical interventions. Discussion about sharing health insights and how to engage biological relatives who may be implicated by an individual’s risk results was also missing. This is not entirely surprising given that these frameworks were not specifically developed with genetic services in mind, but demonstrates that some considerations specific to genetic testing are not addressed by the current literature.

Additionally, implementation frameworks emphasizing health equity have limited guidance for ongoing programs. While the cyclical nature of EquIR (Eslava-Schmalbach et al., 2019) and ongoing evaluation measures provided by RE-AIM (Shelton et al., 2020) can be used to some extent, guidance particularly relevant to genetics again appears to be absent. For instance, frameworks provide little assistance about how to incorporate new risk information over time or ensure that providers are up to date on genetic recommendations so that they can best advise their patients. Again, as the frameworks analyzed here were not designed to be genetics specific, gaps in guidance are expected and suggest a need for frameworks tailored for genetic programs.

### Population genetic screening health equity considerations

Synthesizing findings from the included frameworks, we have compiled a list of relevant health equity questions and outcomes that warrant consideration during the implementation of population genetic screening programs in order to limit health disparities (Table 4). Though not exhaustive, questions may be useful throughout the design and implementation of future screening programs and spur further discussion related to pursing health equity. Broadly, considerations include the accessibility and cultural sensitivity of different population screening processes. Outcomes focus on understanding the distribution of benefits and harms from genetic screening, and the acceptability of program procedures across various demographic groups.

**Table 4:** Health equity considerations for population genetic screening programs.

| Stage | Health equity-focused questions | Outcomes assessed across social determinants |
| --- | --- | --- |
| Recruitment | • If recruitment occurs in-person, is it at an accessible location? Do people have adequate transportation to the site? Are these physical spaces accessible to people with disabilities, including movement, hearing, vision, etc.?<br>• If recruitment occurs online, how can people without regular internet access be reached?<br>• What are relevant cultural beliefs about genetics in specific population groups? Are recruitment materials designed with these in mind?<br>• What language are informational and consent materials provided in? Does this align with people's preferred language?<br>• How does socioeconomic status and insurance coverage influence screening enrollment?<br>• How does a history of harms influence screening enrollment?<br>• If screening is offered by providers, is it offered equally? What provider or patient factors influence if screening is offered? | • Number of people offered screening<br>• Number of people who agree to screening<br>• How do people (e.g., health professionals, community members) perceive screening?<br>• Are recruitment and outreach procedures considered acceptable?<br>• Does pre-screening information lead to informed decision-making about screening? |
| Sample collection | • If sample collection takes place in-person, is it at an accessible location? Do people have adequate transportation to the site? Are these physical spaces accessible to people with disabilities, including movement, hearing, vision, etc.? Does collection take place during routine care?<br>• If sample collection occurs at home, do | • Proportion of people who provide a sample among those who want to receive screening<br>• How do people (e.g., health professionals, community members, stakeholders) perceive |
|  | people have a regular address a collection kit can be sent to and a mailbox for return?<br>• What are relevant cultural beliefs about genetics in specific population groups? Are sample collection and retention procedures designed with these in mind?<br>• What language are materials about sample collection procedures provided in? Does this align with people's preferred language? | the sample collection process? Are procedures considered acceptable?<br>• How easy was it for people to collect samples? If needed, how easy was sample recollection? |
| Return of results | • If return of results occurs in-person, is it at an accessible location? Do people have adequate transportation to the site? Are these physical spaces accessible to people with disabilities, including movement, hearing, vision, etc.?<br>• If return of results occurs online or via phone, how can people without regular internet or phone access be reached?<br>• What are relevant cultural beliefs about genetics in specific population groups? Are clinical services provided with these in mind?<br>• What language are clinical services provided in? Does this align with people's preferred language?<br>• Do all people with the same screening results receive the most appropriate level of guidance? | • Proportion of people who receive results among those who provide samples<br>• Proportion of people who indicate experiencing psychosocial harms among those who receive screening results<br>• How do people perceive the return of results process? Is the guidance provided acceptable?<br>• How helpful or useful do people find the information learned through screening?<br>• How much time is present between when people provide samples and when results are returned? |
| Follow-up care & prevention | • Are necessary clinics or specialists in | • Proportion of people |
|  | accessible locations? Do people have adequate transportation to relevant facilities? Are these physical spaces accessible to people with disabilities, including movement, hearing, vision, etc.?<br>• How does socioeconomic status and insurance coverage influence prevention uptake?<br>• Are all people with the same risk profiles referred to the same type of specialists or advised in the same way? | who discuss results with their provider among those receiving screening results^<br>• Proportion of people who meet with appropriate specialists among those who receive positive risk results<br>• Proportion of people who adhere to appropriate medical interventions among those who receive positive risk results<br>• Proportion of people who experience psychosocial harms or clinical harms^ |
| Cascade screening | • Are genetic services accessible to biological relatives?<br>• How do health beliefs, health literacy, and family dynamics influence how genetic risk is discussed within families?<br>• Are all individuals offered the same support regarding risk communication?<br>• What are local/state considerations for cascade screening (e.g., related to sharing genetic information)? | • Proportion of people who discuss genetic risk with biological relatives among those receiving screening results^<br>• Number of biological relatives who receive testing<br>• How do people view genetic risk information sharing? Is such sharing considered acceptable? |
| Overall considerations | • Are community partners and other stakeholders involved in program planning, design implementation, and evaluation?<br>• What processes are in place to facilitate program adaptations? | • To what degree do community partners or stakeholders report understanding of and involvement in program processes, trust in research partners, or benefits from program implementation?<br>• How often are program procedures reviewed? By whom are they reviewed?<br>• After receiving screening, would people recommend screening to others? |

^Consider by type of screening result (e.g., positive or uninformative)

One of the overall considerations for pursuing health equity is involving community partners (Table 4). As members of the community are likely more in-tune with local settings compared to researchers, they may be better equipped to understand and identify drivers behind complex inequities (*Leveraging Community Expertise to Advance Health Equity*, 2021). Through community engagement, researchers and public health professionals can ascertain what communities identify as problems to be addressed and what community health priorities are. This can inform if population genetic screening is a suitable intervention in a particular setting and truly meeting community needs. Investment by communities in population genetic screening programs can also promote sustainability of such programs.

Even with these health equity considerations identified, challenges may emerge when incorporating these ideas into practice. For example, answers to these questions may vary by communities included in a single screening program. Resource constraints may also prevent the adoption of more equitable practices. Additionally, outcome measures may be difficult to ascertain if they involve time-intensive data collection and the continued engagement of people who have taken part in genetic screening. As such, researchers and health professionals looking to implement screening programs may benefit from using these considerations to appropriately plan and allocate resources.

### Limitations

Our study may have been limited by the frameworks considered for analysis. Due to stringent inclusion criteria, we may not have identified all relevant frameworks. In addition, though we give a broad overview of implementation science frameworks that center health equity, we did not assess the quality of these frameworks. However, our analysis of how frameworks have been used in other settings provides an indirect measure of utility and quality. Our assessment of framework applicability to population genetic screening programs may also have been limited. Relevant stages, such as sample lab testing, were not included and some of our analysis may be applicable to multiple stages though not detailed here. Despite these limitations, our study is a first step in describing the current state of implementation science frameworks that explicitly focus on health equity and how they can be applied to improve the equitable implementation of population genetic screening programs.

## CONCLUSION

Current implementation science frameworks that emphasize health equity offer broad recommendations applicable to the implementation of population genetic screening programs. However, gaps still exist in guidance provided for stages of screening that are ongoing, such as follow-up care and cascade screening. Through our application of frameworks to population genetic screening, we have created a list of considerations and outcomes that may assist with more equitable implementation. Researchers planning to implement screening programs may benefit from consulting these considerations or following guidance from analyzed frameworks.

## Data Availability

All data produced in the present work are contained in the manuscript

## REFERENCES

Baumann, A. A., & Cabassa, L. J. (2020). Reframing implementation science to address inequities in healthcare delivery. BMC Health Services Research, 20(1), 190. 10.1186/s12913-020-4975-3

Braveman, P. (2014). What are Health Disparities and Health Equity? We Need to Be Clear. Public Health Reports, 129(1_suppl2), 5–8. 10.1177/00333549141291S203

Cabassa, L. J., & Baumann, A. A. (2013). A two-way street: Bridging implementation science and cultural adaptations of mental health treatments. Implementation Science, 8(1), 90. 10.1186/1748-5908-8-90

Centers for Disease Control and Prevention. (n.d.). What is Health Equity? Retrieved July 15, 2022, from https://www.cdc.gov/nchhstp/healthequity/index.html

Chambers, D. A., Feero, W. G., & Khoury, M. J. (2016). Convergence of Implementation Science, Precision Medicine, and the Learning Health Care System: A New Model for Biomedical Research. JAMA, 315(18), 1941–1942. 10.1001/jama.2016.3867

David, S. P., Dunnenberger, H. M., Ali, R., Matsil, A., Lemke, A. A., Singh, L., Zimmer, A., & Hulick, P. J. (2021). Implementing Primary Care Mediated Population Genetic Screening Within an Integrated Health System. The Journal of the American Board of Family Medicine, 34(4), 861–865. 10.3122/jabfm.2021.04.200381

Drake, C., Batchelder, H., Lian, T., Cannady, M., Weinberger, M., Eisenson, H., Esmaili, E., Lewinski, A., Zullig, L. L., Haley, A., Edelman, D., & Shea, C. M. (2021). Implementation of social needs screening in primary care: A qualitative study using the health equity implementation framework. BMC Health Services Research, 21(1), 975. 10.1186/s12913-021-06991-3

East, K. M., Kelley, W. V., Cannon, A., Cochran, M. E., Moss, I. P., May, T., Nakano-Okuno, M., Sodeke, S. O., Edberg, J. C., Cimino, J. J., Fouad, M., Curry, W. A., Hurst, A. C. E., Bowling, K. M., Thompson, M. L., Bebin, E. M., Johnson, R. D., AGHI Consortium, Cooper, G. M., … Korf, B. R. (2021). A state-based approach to genomics for rare disease and population screening. Genetics in Medicine: Official Journal of the American College of Medical Genetics, 23(4), 777–781. 10.1038/s41436-020-01034-4

Eslava-Schmalbach, J., Garzón-Orjuela, N., Elias, V., Reveiz, L., Tran, N., & Langlois, E. V. (2019). Conceptual framework of equity-focused implementation research for health programs (EquIR). International Journal for Equity in Health, 18(1), 80. 10.1186/s12939-019-0984-4

Glasgow, R. E., Harden, S. M., Gaglio, B., Rabin, B., Smith, M. L., Porter, G. C., Ory, M. G., & Estabrooks, P. A. (2019). RE-AIM Planning and Evaluation Framework: Adapting to New Science and Practice With a 20-Year Review. Frontiers in Public Health, 7. https://www.frontiersin.org/articles/10.3389/fpubh.2019.00064

Glasgow, R. E., Vogt, T. M., & Boles, S. M. (1999). Evaluating the public health impact of health promotion interventions: The RE-AIM framework. American Journal of Public Health, 89(9), 1322–1327.

Grzymski, J. J., Coppes, M. J., Metcalf, J., Galanopoulos, C., Rowan, C., Henderson, M., Read, R., Reed, H., Lipp, B., Miceli, D., Rybarski, S., & Slonim, A. (2018). The Healthy Nevada Project: Rapid recruitment for population health study (p. 250274). bioRxiv. 10.1101/250274

Grzymski, J. J., Elhanan, G., Morales Rosado, J. A., Smith, E., Schlauch, K. A., Read, R., Rowan, C., Slotnick, N., Dabe, S., Metcalf, W. J., Lipp, B., Reed, H., Sharma, L., Levin, E., Kao, J., Rashkin, M., Bowes, J., Dunaway, K., Slonim, A., … Lu, J. T. (2020). Population genetic screening efficiently identifies carriers of autosomal dominant diseases. Nature Medicine, 26(8), Article 8. 10.1038/s41591-020-0982-5

Harvey, G., & Kitson, A. (2016). PARIHS revisited: From heuristic to integrated framework for the successful implementation of knowledge into practice. Implementation Science, 11(1), 33. 10.1186/s13012-016-0398-2

Kilbourne, A. M., Switzer, G., Hyman, K., Crowley-Matoka, M., & Fine, M. J. (2006). Advancing Health Disparities Research Within the Health Care System: A Conceptual Framework. American Journal of Public Health, 96(12), 2113–2121. 10.2105/AJPH.2005.077628

King, M.-C., Levy-Lahad, E., & Lahad, A. (2014). Population-based screening for BRCA1 and BRCA2: 2014 Lasker Award. JAMA, 312(11), 1091–1092. 10.1001/jama.2014.12483

Leveraging Community Expertise to Advance Health Equity. (2021, July 1). Urban Institute. https://www.urban.org/research/publication/leveraging-community-expertise-advance-health-equity

Limburg, P. J., Harmsen, W. S., Chen, H. H., Gallinger, S., Haile, R. W., Baron, J. A., Casey, G., Woods, M. O., Thibodeau, S. N., & Lindor, N. M. (2011). Prevalence of alterations in DNA mismatch repair genes in patients with young-onset colorectal cancer. Clinical Gastroenterology and Hepatology: The Official Clinical Practice Journal of the American Gastroenterological Association, 9(6),497–502. 10.1016/j.cgh.2010.10.021

Marquez, C., Kerkhoff, A. D., Naso, J., Contreras, M. G., Diaz, E. C., Rojas, S., Peng, J., Rubio, L., Jones, D., Jacobo, J., Rojas, S., Gonzalez, R., Fuchs, J. D., Black, D., Ribeiro, S., Nossokoff, J., Tulier-Laiwa, V., Martinez, J., Chamie, G., … Havlir, D. V. (2021). A multi-component, community-based strategy to facilitate COVID-19 vaccine uptake among Latinx populations: From theory to practice. PLOS ONE, 16(9), e0257111. 10.1371/journal.pone.0257111

Nápoles, A. M., & Stewart, A. L. (2018). Transcreation: An implementation science framework for community-engaged behavioral interventions to reduce health disparities. BMC Health Services Research, 18(1), 710. 10.1186/s12913-018-3521-z

Nilsen, P. (2015). Making sense of implementation theories, models and frameworks. Implementation Science, 10(1), 53. 10.1186/s13012-015-0242-0

Odeny, B. (2021). Closing the health equity gap: A role for implementation science? PLOS Medicine, 18(9), e1003762. 10.1371/journal.pmed.1003762

Proctor, E. K., Landsverk, J., Aarons, G., Chambers, D., Glisson, C., & Mittman, B. (2009). Implementation Research in Mental Health Services: An Emerging Science with Conceptual, Methodological, and Training challenges. Administration and Policy in Mental Health and Mental Health Services Research, 36(1), 24–34. 10.1007/s10488-008-0197-4

Rao, N. D., Kaganovsky, J., Malouf, E. A., Coe, S., Huey, J., Tsinajinne, D., Hassan, S., King, K. M., Fullerton, S. M., Chen, A. T., & Shirts, B. H. (2023). Diagnostic yield of genetic screening in a diverse, community-ascertained cohort. Genome Medicine, 15(1), 26. 10.1186/s13073-023-01174-7

Roberts, M. C., Kennedy, A. E., Chambers, D. A., & Khoury, M. J. (2017). The current state of implementation science in genomic medicine: Opportunities for improvement. Genetics in Medicine, 19(8), 858–863. 10.1038/gim.2016.210

Santoyo-Olsson, J., Stewart, A. L., Samayoa, C., Palomino, H., Urias, A., Gonzalez, N., Torres-Nguyen, A., Coleman, L., Escalera, C., Totten, V. Y., Ortiz, C., & Nápoles, A. M. (2019). Translating a stress management intervention for rural Latina breast cancer survivors: The Nuevo Amanecer-II. PLOS ONE, 14(10), e0224068. 10.1371/journal.pone.0224068

Shelton, R. C., Chambers, D. A., & Glasgow, R. E. (2020). An Extension of RE-AIM to Enhance Sustainability: Addressing Dynamic Context and Promoting Health Equity Over Time. Frontiers in Public Health, 8. https://www.frontiersin.org/articles/10.3389/fpubh.2020.00134

Wallerstein, N., & Duran, B. (2010). Community-Based Participatory Research Contributions to Intervention Research: The Intersection of Science and Practice to Improve Health Equity. American Journal of Public Health, 100(S1), S40–S46. 10.2105/AJPH.2009.184036

Woodward, E. N., Matthieu, M. M., Uchendu, U. S., Rogal, S., & Kirchner, J. E. (2019). The health equity implementation framework: Proposal and preliminary study of hepatitis C virus treatment. Implementation Science, 14(1), 26. 10.1186/s13012-019-0861-y

Woodward, E. N., Singh, R. S., Ndebele-Ngwenya, P., Melgar Castillo, A., Dickson, K. S., & Kirchner, J. E. (2021). A more practical guide to incorporating health equity domains in implementation determinant frameworks. Implementation Science Communications, 2(1), 61. 10.1186/s43058-021-00146-5

